# VS-FPM: large-format, label-free virtual histopathology microscopy

**DOI:** 10.1101/2025.05.20.25327933

**Authors:** Christopher Bendkowski, Adam P. Levine, Manuel Rodriguez-Justo, Laurence B. Lovat, Marco Novelli, Michael Shaw

## Abstract

By generating realistic histologically-stained images from label-free image data, virtual staining (VS) methods have the potential to streamline clinical workflows, improve image consistency and provide new ways of visualizing and analysing tissues. This article describes a new VS approach based on the application of conditional generative adversarial networks to translate high-resolution phase images of unstained tissues, recovered using Fourier ptychographic microscopy (FPM), into brightfield H&E images. Compared to other label-free imaging methods, FPM offers unique advantages for VS as it allows simultaneous capture of sample amplitude and phase information, simplifying the pixelwise registration required for supervised training. FPM combines high spatial resolution with a large field of view and a large depth of field making it well suited to large format imaging of histological tissues. The method is readily implemented by modifying a conventional brightfield microscope using simple, low-cost optoelectronic hardware. Using colonic polyps as a test case, we compare FPM and VS-FPM images to brightfield whole slide images (WSIs) captured using a pathology slide scanner. Our results show that FPM images captured at 4x magnification have a spatial resolution equivalent to WSIs captured at 20x magnification. Virtual H&E images of unstained tissues generated using VS-FPM closely match brightfield images of the same tissue sections captured after chemical staining, enabling pathological assessment and diagnosis.

**Highlights:**

- Fourier Ptychography (FPM) can capture large-format complex images of tissue sections.
- 4x FPM images and 20x images of H&E-stained tissues have equivalent spatial resolution.
- Conditional GANs can generate accurate brightfield H&E images from FPM phase images.
- VS-FPM enabled pathologists to assess and diagnose unstained colonic polyps.
- The method can be applied to other sample types in histo- and cyto-pathology.

**Graphical abstract:** 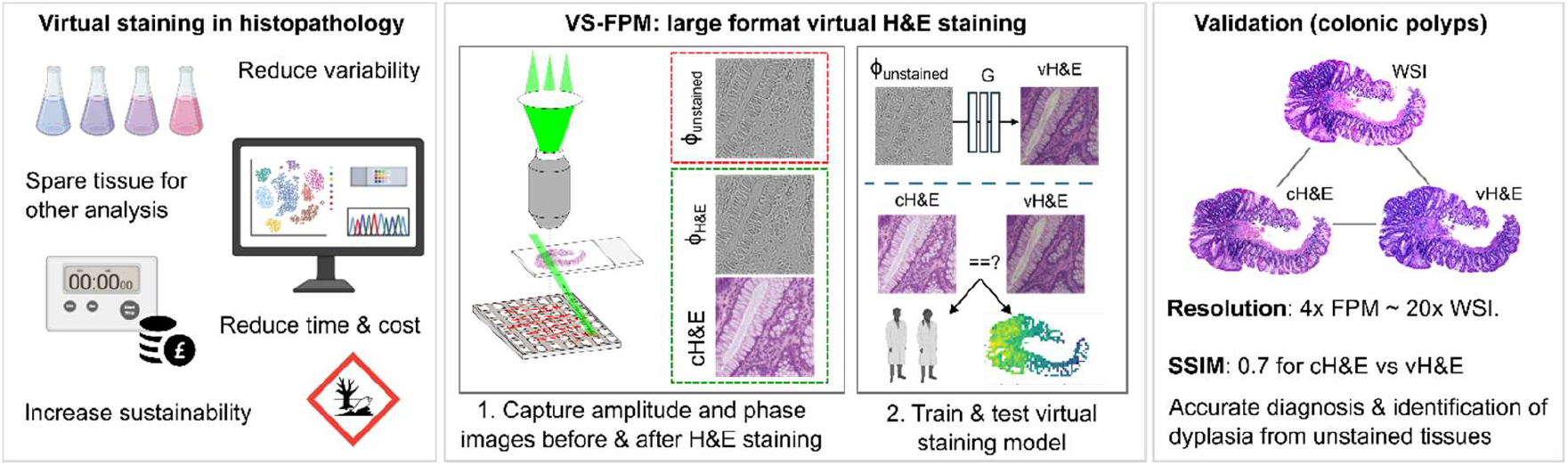

## 1. Introduction

Conventional histopathology is based on the analysis of thin sections of stained tissues using brightfield microscopy. Chemical stains enable visualization of tissue structure, cellular and subcellular organization and morphology, and the distribution of specific proteins for diagnosis, prognosis and assessment of the efficacy of therapeutic interventions. However, chemical staining adds time to clinical workflows, increases costs, and results in toxic byproducts and significant amounts of wastewater. Differences between reagents and staining protocols increases intra-and inter-laboratory variability, which is of growing importance with the increasing utilisation of artificial intelligence based-image analysis (Bahadir et al., 2024). Extensive tissue processing increases the risk of specimen contamination (carry over). Typically, a single stain is applied to each tissue section and once stained the same piece of tissue is often not suitable for further assessment using complementary biochemical and molecular analysis. As a result, virtual staining (VS) methods, in which a deep neural network (DNN) is trained to transform label-free image data into an equivalent histologically-stained brightfield image, present an attractive alternative to the use of chemical stains in both diagnostic histopathology and research (Bai et al., 2023).

Since the DNN-based virtual staining concept was first introduced (Rivenson et al., 2019b) a number of different imaging techniques have been employed for generating contrast from unstained tissues, including autofluorescence (Rivenson et al., 2019b) and fluorescence lifetime microscopy (Wang et al., 2024), quantitative phase imaging (Rivenson et al., 2019a), and photoacoustic microscopy and reflected light confocal microscopy (Li et al., 2024). A variety of supervised and unsupervised DNN architectures have been used for the task of translating label-free images into the histologically-stained brightfield images required for diagnostic assessment by pathologists (Bai et al., 2023). Generative adversarial networks (GANs), including conditional and cycle GANs are most commonly employed, however other methods such as diffusion models have also been proposed (Zhang et al., 2024).

In this article we introduce a new method (VS-FPM) for virtual histological staining based on the application of Fourier ptychographic microscopy (FPM) to capture complex images of unstained slide-mounted tissue. High-resolution FPM phase images are virtually-stained using a DNN trained in a conditional GAN framework using paired FPM phase and amplitude images captured before and after chemical H&E staining. Compared to other label-free imaging methods, FPM offers significant advantages for digital pathology (Horstmeyer et al., 2015), including experimental simplicity and low hardware cost. By reconstructing high-resolution images from data captured using a low magnification (low numerical aperture) objective lens, FPM enables large format, high-information content imaging of large tissue sections without stage scanning. Recovery of sample amplitude and phase allows images to be digitally refocused after capture (Claveau et al., 2020) for correction of sample tilt and focus errors. Complex images captured before and after chemical staining significantly simplify the pixelwise registration of image data for DNN training compared to other VS methods

To test VS-FPM we built a simple FPM platform using off-the-shelf components and captured images of formalin fixed paraffin embedded (FFPE) sections of colonic polyp biopsies before and after H&E staining. Comprising a range of different cell types (including epithelial, stromal and inflammatory cells) and dysplastic and normal regions, colonic polyps enabled evaluation of the performance of VS-FPM for different tissue structures and cell morphologies. We compare FPM images of chemically H&E-stained tissues captured using a 4x/0.16 objective lens to images captured using a 20x/0.75 objective lens with a state-of-the-art brightfield slide scanning system, virtually H&E-stained FPM phase images to chemically H&E-stained FPM amplitude images, and virtually H&E-stained FPM phase images to brightfield WSIs of stained tissues; evaluating results using image similarity metrics, empirically estimated spatial resolution and qualitative diagnostic assessment. Our results demonstrate that VS-FPM is an accessible, clinically valuable method for histological examination of unstained tissues. Finally, we discuss the relative advantages and limitations of VS-FPM compared to other VS methods and wider applications of the technique in research and clinical practice.

## 2. Methods

### 2.1 Overview of methods

**Figure 1.**
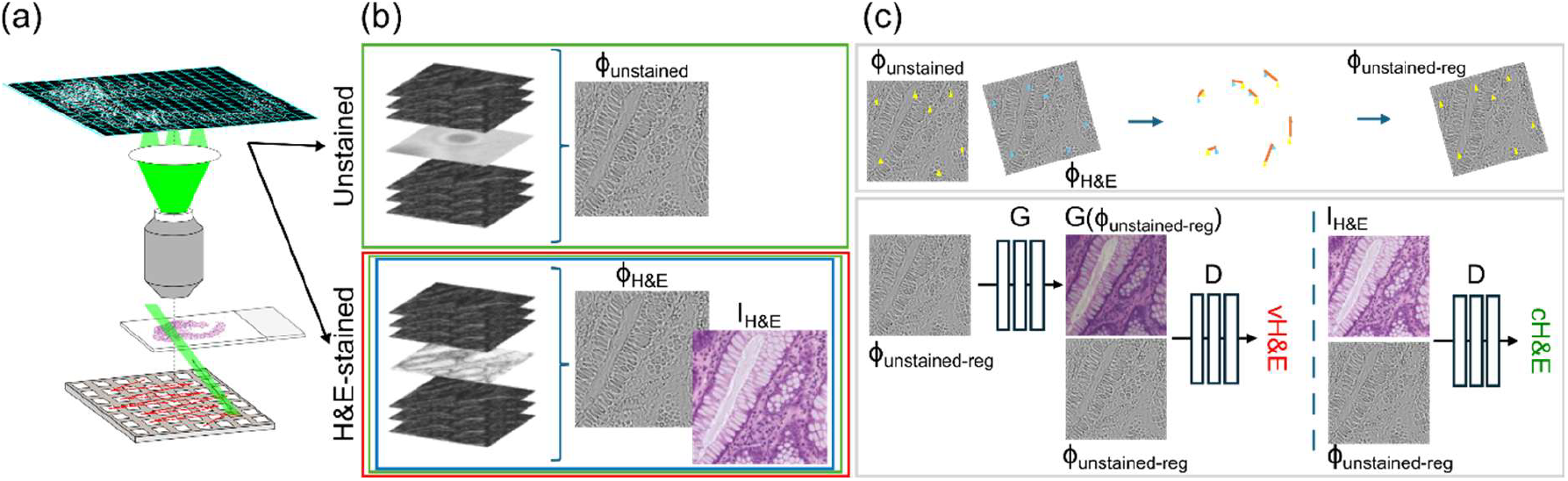
Overview of experimental methods. (a) Schematic diagram of FPM image acquisition system. A sequence of raw images is captured under inclined illumination of the sample using an LED array. (b) Brightfield and darkfield images are processed using iterative phase retrieval to reconstruct high-resolution estimates of the sample phase before chemical staining (top) and the phase and amplitude of the same tissue section after H&E staining (bottom). (c) Phase images of the same tissue section captured before and after chemical H&E staining enable pixelwise registration (top). Pairs of phase and amplitude images captured before and after chemical H&E staining are then used to train a virtual staining model in generative adversarial network (bottom).

### 2.2 Slide preparation

Anonymized FFPE blocks from colonic polyps (tubular adenomata with low grade dysplasia) that were surplus to diagnostic requirements were obtained from a clinical histopathology archive. 4 µm thick sections were cut using a microtome, mounted on standard glass microscope slides and deparaffinized. Each section was then sealed with DPX mountant and a coverslip to minimize contamination. After FPM imaging, coverslips were removed by submerging the slides in xylene for 48 hours before H&E staining and sealing with a new coverslip.

### 2.3 Fourier ptychographic microscopy

Image data were captured using a custom-built upright FPM system (Shaw et al., 2021). In which samples were illuminated individually by (red, green and blue) LEDs in a 22×22 array (7 mm pitch, WS212B, WORLDSEMI) controlled via an ESP32-based microcontroller. A 3D printed conical aperture was mounted above the LED array to minimise stray light caused by shallow angle reflections between the lower surface of the glass slide and the top surface of the LED packages. Each raw, monochromatic, FPM data set comprised 177 images corresponding to sequential illumination from the LEDs within a circle centred on the optical axis. With the LED array at distance of 81 mm below the top surface of the sample slide, this resulted in reconstructed images with a synthetic NA (*NA_syn_*= *NA_obj_*+ sin *θ_max_*, where *NA_obj_*is the NA of the objective lens and *θ_max_*the largest illumination angle with respect of the optic axis) of approximately 0.7. The imaging pathway consisted of a 4x/0.16 objective lens (UPLSAPO, Olympus) mounted on a piezoelectric focus scanner (P-725.1CDE2, Physik Instrumente) and a tube lens with a focal length of 200 mm (TTL200-A, Thorlabs Inc.), resulting in an effective magnification of 4.44. Images were captured using a large format scientific CMOS camera (IRIS 15, Photometrics) synchronized to the LED array. This setup provided a field of view (FoV) of 4.8 mm x 2.8 mm – in many cases sufficient to capture an entire polyp section within a single image. The focus scanner was used for autofocusing at each wavelength before image acquisition by maximising the normalised variance of images captured under darkfield illumination from a ring of LEDs on the LED array. Microscope slides were mounted on a motorised XY stage (H101A, Prior Scientific) for sample positioning and coarse registration of images captured before and after H&E staining. Images of unstained tissue sections were captured under illumination by green LEDs (CWL ∼ 530 nm) as a compromise between FoV, lateral resolution, image contrast and sampling in Fourier space. Colour images of H&E-stained tissue sections were captured under sequential illumination by red (CWL ∼ 629 nm), green and blue (CWL ∼ 475 nm) LEDs. Brightfield images were captured with an exposure time (*T_BF_*) of 2 ms. To improve the signal-to-noise ratio the exposure time (*T_n_*) for each darkfield image was adjusted in proportion to the squared distance *d_n_*of the illuminating LED from the optical axis, *T_n_*= *T_BF_*· *d*_*n*_^2^. The intensity *I* of the captured images was then scaled down accordingly, *I_n_*= *I_raw_*· *T_n_*/*T_BF_*.

Captured FPM sequences (177 5056 x 2960 pixel images) were processed using an iterative Gauss-Newton phase retrieval algorithm (Yeh et al., 2015) to reconstruct high-resolution complex images. For reliability and computational simplicity, images were reconstructed in patches of 243 x 243 pixels and upsampled by a factor of three to accommodate the additional high-resolution information. Scalar offsets and gradients between phase image patches were corrected by normalising each patch by first subtracting the mean phase and setting the standard deviation to unity. A Gaussian blur (with a kernel size 31 pixels) was then applied to a copy of the patch and this blurred patch was subtracted from the normalised patch. Background regions in the corrected phase images were thereby set to ∼0.

To correct for intensity variations in reconstructed amplitude images of chemically-stained sections, the high-resolution amplitude patch and the corresponding brightfield low-resolution patch (formed from the sum of the brightfield images captured in the FPM stack) were histogram matched. Following reconstruction and correction, amplitude and phase patches were stitched using the Grid/Collection stitching ImageJ plugin (Preibisch et al., 2009) to reform a full high-resolution FoV (15142 x 8867 pixels). Radial and tangential distortion in the high-resolution images were then corrected using calibrated camera parameters to give the flat image necessary for image registration before and after staining. To correct for lateral chromatic offsets, red, green and blue amplitude images of H&E-stained sections were aligned using an affine transform derived from matched SIFT (Lowe, 2004) features. The aligned RGB images were then self white balanced using a manually-defined background region of interest (RoI).

### 2.4 Whole slide imaging

WSIs of H&E-stained polyps were captured using a NanoZoomer S360MD (Hamamatsu Photonics K. K.) in 40x mode with a pixel size of 230 nm. The images were stored in a pyramidal format and the (downsampled) 20x images were used for comparison to FPM image results for stained and unstained sections.

### 2.5 Virtual H&E staining

Training of the virtual staining network used pixelwise registered pairs of FPM phase images of unstained tissues and the corresponding RGB FPM amplitude images after H&E staining. Absorption of light by H&E modified the local refractive index of the stained tissue sections and had the effect of blurring out some of the fine features visible in phase images of unstained sections (see supplementary Fig.S1). However, the overall similarity of FPM phase images before and after chemical staining enabled reliable image registration of complex images. Matched SIFT features (Lowe, 2004) were used to find corresponding keypoints in the unstained and stained phase images, before RANSAC (Fischler and Bolles, 1987) was applied to find a perspective transform. As the virtual staining network required small patches to train on, the high-resolution matched sets of unstained phase and stained phase and amplitude images were divided into patches of 256 x 256 pixels. Each patch was then assessed using mutual information of the phase image histograms to determine if the match was of sufficient quality to be included in the training dataset. A cutoff value of 0.4 was used to exclude outlier patches, which included those containing reconstruction artefacts or those corresponding to tissue-free background parts of the slide.

Virtual staining was performed using the Pix2Pix cGAN architecture (Isola et al., 2017) with an objective defined as

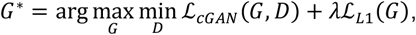

where *ℒ_cGAN_*(*G, D*) is the objective of the cGAN as defined in the original Pix2Pix article (Isola et al., 2017), *λℒ*_*L*1_(*G*) is the L1 loss of the generator and *λ* = 100. The training phase comprised 100 epochs and used a dataset of 17199 matched pairs of 256 x 256 pixel unstained phase and H&E-stained RGB amplitude images sampled from 12 different tissue sections. The dataset was augmented during training by applying random jitter and mirroring. During inference, the memory limitations of the GPU (RTX 3090, NVIDIA) restricted the image size that could be virtually stained, so the high-resolution (15142 x 8867 pixels) FoVs were divided into 1024 x 1024 pixel patches which were then stitched using the Grid/Collection stitching ImageJ plugin (Preibisch et al., 2009) to give a virtually-stained FoV. When required, FoVs were stitched using the BigStitcher ImageJ plugin (Hörl et al., 2019) to produce an image of a whole slide.

### 2.6 Image evaluation

Differences between brightfield WSIs, FPM amplitude images of H&E-stained tissues, and VS-FPM images unstained tissues were quantified using structural similarity index (SSIM), peak signal to noise ratio (PSNR) and root mean square error (RMSE). For a pair of images *x* and *y*, each comprising *M* · *N* pixels and with a maximum pixel value of *R*, these were defined as,

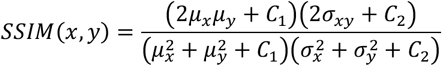

Where *μ_*x*_*and *σ*_*x*_are the mean and standard deviation of image *x, σ_xy_*is the covariance of images *x* and *y* and *C*_1_ and *C*_2_ are small constants used to avoid instability.

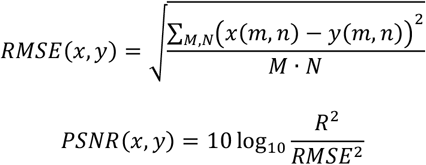

Image resolution was quantified using the spatial frequency cutoff estimated using image decorrelation analysis, as described in (Descloux et al., 2019). Briefly, a decorrelation function was computed from the cross correlation of the 2D Fourier transform (FT) of the image normalised by its amplitude and filtered by a series of circular binary masks with the unnormalized, unmasked 2D FT of the image. Decorrelation functions were then computed for a series of high pass filtered images and the cutoff was defined as the highest frequency at which a local maximum was detected in the decorrelation function.

## 3. Results

### 3.1 Comparison of FPM and WSIs of chemically H&E-stained sections

To compare the imaging performance of FPM (with a 4x/0.16 objective lens) to a brightfield slide scanner (with a 20x/0.75 objective lens) we captured and analysed images of the same set of 18 H&E-stained polyp sections using both systems. The large FoV of the FPM system meant that a polyp section typically fit within the FPM field of view, minimising the need for stage scanning. Figure 2(a) shows representative low magnification overview images of one section from both imaging systems. The shape of the tissue section, the arrangement and morphology of large features such as crypts, and variations in the number density of cell nuclei are clearly visible in both the FPM and WSI results. The magnified RoIs (Fig. 2(b)) illustrate that detailed features, including the spatial organization of the tissue, cellular morphology, variations in haematoxylin staining and the detailed structures of crypts, are also clearly resolved in the FPM images. The most striking visual difference between the WSI and FPM results is the colour of the images. FPM colour images were generated from three monochrome images captured sequentially under narrowband illumination from red, green and blue LEDs, whereas WSIs were captured using a slide scanner with a colour camera and white light illumination. As a result, the imaging systems are expected have very different colour gamuts (Cheng, 2020) and simple self-white balancing is insufficient to match the colour of the two images. The effect of improved colour correction/white balancing on the visual appearance of the FPM is highlighted by the lower row of magnified images (Fig. 2(b)) in which the reconstructed FPM amplitude image was histogram matched to the WSI.

**Figure 2.**
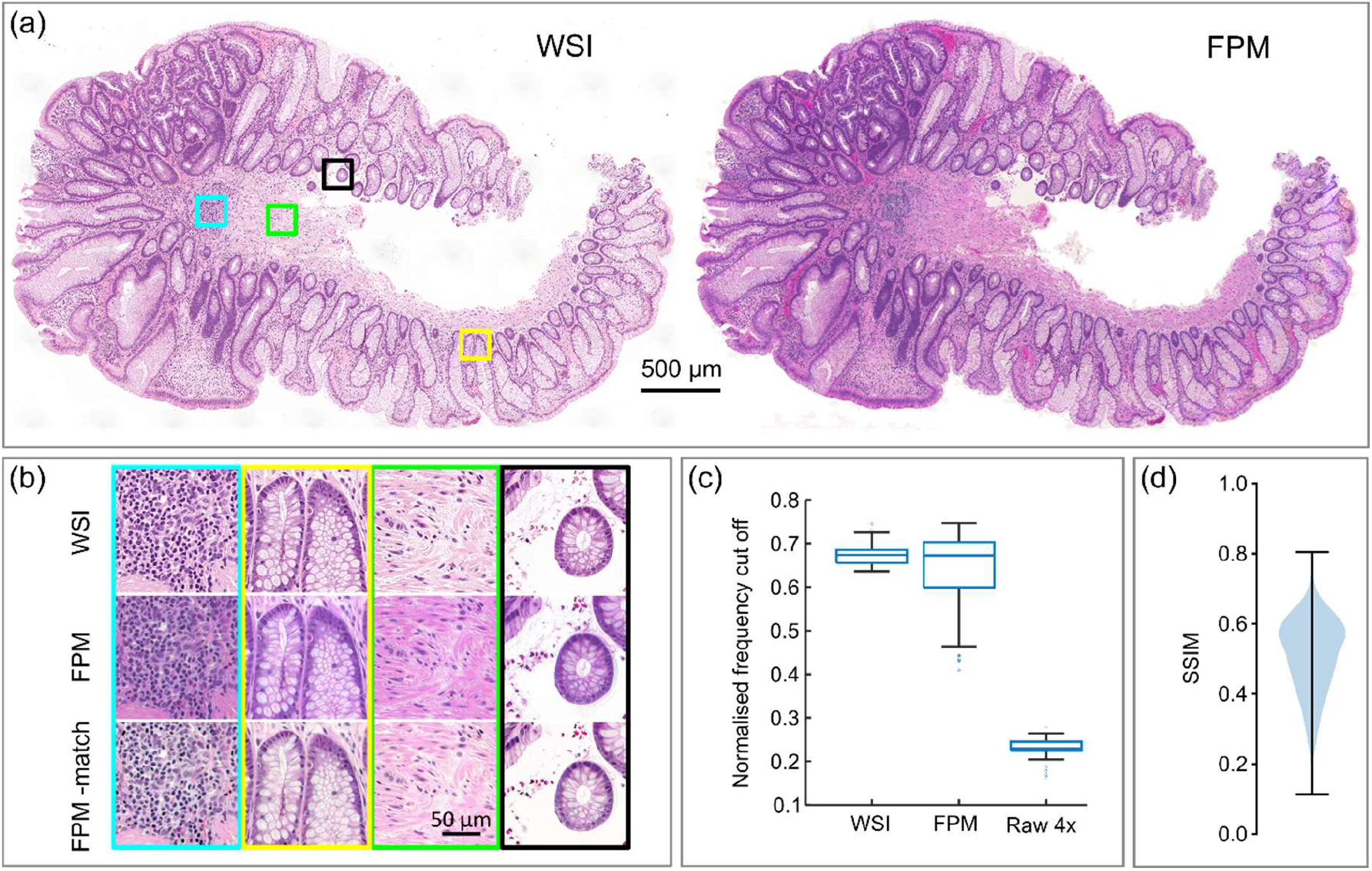
Comparison of WSIs and FPM images of H&E-stained colonic polyps. (a) Representative WSI and FPM amplitude images of an H&E-stained polyp section. (b) Magnified images of the boxed regions shown in panel (a), showing WSI (top row), FPM image with simple self-white balancing (middle row) and FPM image histogram matched to the WSI (bottom row). (c) Box plots showing the empirical cut of frequency for WSI, FPM and raw 4x images of the section shown in (a) estimated using decorrelation analysis. (d) Violin plot of structural similarity index between WSI and FPM amplitude images from 18 different polyp sections.

The nominal spatial resolution of a reconstructed FPM image is typically defined by the nominal synthetic numerical aperture of the system (*NA_syn_*), with the cutoff frequency given by *f_max_*= *λ*/*NA_syn_*. The NA of the objective lens in the slide scanner was 0.75, similar to the nominal *NA_syn_*of 0.7 of the FPM hardware; as such images from both systems were expected to have similar spatial resolution. However, in practice the presence of imaging aberrations and noise affect the imaging performance and resolving power. To empirically compare the lateral spatial resolution of the WSIs and FPM images we used decorrelation analysis to estimate the spatial frequency cutoff for 90 512×512 pixel patches from the images shown in Fig. 2(a). Analysis of the results using Welch’s t-test indicated no statistically significant difference (p = 0.29) between the mean cutoff frequency for the WSI and FPM image patches - 0.67 and 0.68 respectively. However, as the cutoff frequency box plots (Fig. 2(c)) indicate, there was a significantly greater variance (0.095) in measured cutoff frequency for the FPM images than for the WSI results (0.024). In FPM the amount of light scattered into the objective lens at darkfield illumination angles is strongly dependent on the local sample properties - in particular the magnitude of refractive index differences due to the inhomogeneity of the structure and physico-chemical composition of the tissue section. Thus we hypothesize that the greater variance in measured cutoff is due, at least in part, to differences in the amount of information carried by the darkfield images for each image patch. This is supported by the spatial frequency cut off heatmaps in Fig. S2, which show the frequency cutoff is higher for FPM image patches close to the edges of polyp sections which are spatially inhomogeneous and contain dysplastic crypts characterized by densely packed epithelial cells. Closer to the section centre, where cells are sparse and there are fewer crypts, the cut off frequency is significantly lower. To quantify the resolution increase from a 4x image to the high-resolution FPM reconstructed amplitude image, the spatial frequency cutoff was also estimated for the same 90 patches in the raw (4x) brightfield image (computed as a linear sum over all 177 images in the raw FPM image sequence). The mean cutoff frequency for the 4x brightfield images is 0.26, indicating that the spatial resolution of the reconstructed amplitude is approximately 2.6 times greater that of raw image. As with the WSI images, and consistent with the hypothesis than the spatial resolution in the FPM reconstructions varies with the information present in the darkfield images, the variance in the cutoff estimate is significantly lower for the raw 4x brightfield images than for the reconstructed FPM images.

The box plot in Fig. 2(d) shows the structural similarity index (SSIM) values between the WSI and FPM amplitude images measured for 20,597 patches of 256x256 pixels from all 18 H&E-stained polyp sections. The results (mean SSIM of 0.51 ± 0.11) reflect the differences in colour and spatial resolution discussed above as well as WSI and FPM image acquisition (focusing) and reconstruction (stitching and blending) errors. Heatmaps (Fig. S3(a)) highlight that SSIM values tend to vary gradually over a tissue section, suggesting that sample tilt, thickness variation or mounting errors may explain some of the differences. There is no widely accepted SSIM threshold in digital pathology, however a study of colour normalization techniques for histological images (Vahadane et al., 2016) found SSIM values of between 0.7-0.8 for two, conceptually very similar, commercial WSI systems.

### 3.2 Comparison of chemically H&E-stained FPM amplitude and virtually H&E-stained FPM phase images

Having established that FPM amplitude images of H&E-stained polyp sections captured using a 4x objective are similar to brightfield WSI images captured at 20x we next investigated the accuracy with which FPM phase images of unstained tissues could be virtually H&E stained using a conditional generative adversarial network. After training using pixelwise registered pairs of FPM phase and amplitude patches from 12 polyp sections captured before and after H&E staining, the generator was used to virtually stain reconstructed FPM phase images of six unstained sections excluded from the training set. Figure 3(a) shows representative low magnification images of one of these sections. There is a close visual resemblance between the FPM amplitude image of the H&E-stained section (cH&E image) and the VS-FPM (vH&E image). Viewed at high magnification (Fig. 3(b)) vH&E images contain much of the detailed information visible in the cH&E images. Significantly, normal and dyplastic crypts, characterised by differences in morphology and the number and density of cells and nuclei, can be clearly distinguished in both cH&E and vH&E images. We anticipated that the downsampling half of the U-Net architecture of the VS model would result in a reduction in spatial resolution. As previously, this resolution loss was quantified using decorrelation analysis to estimate the spatial frequency cut off in corresponding cH&E and vH&E FPM images, with the result (Fig. 3(c)) that the mean cut off frequency in the vH&E images was found to be 79% of that in cH&E images.

**Figure 3.**
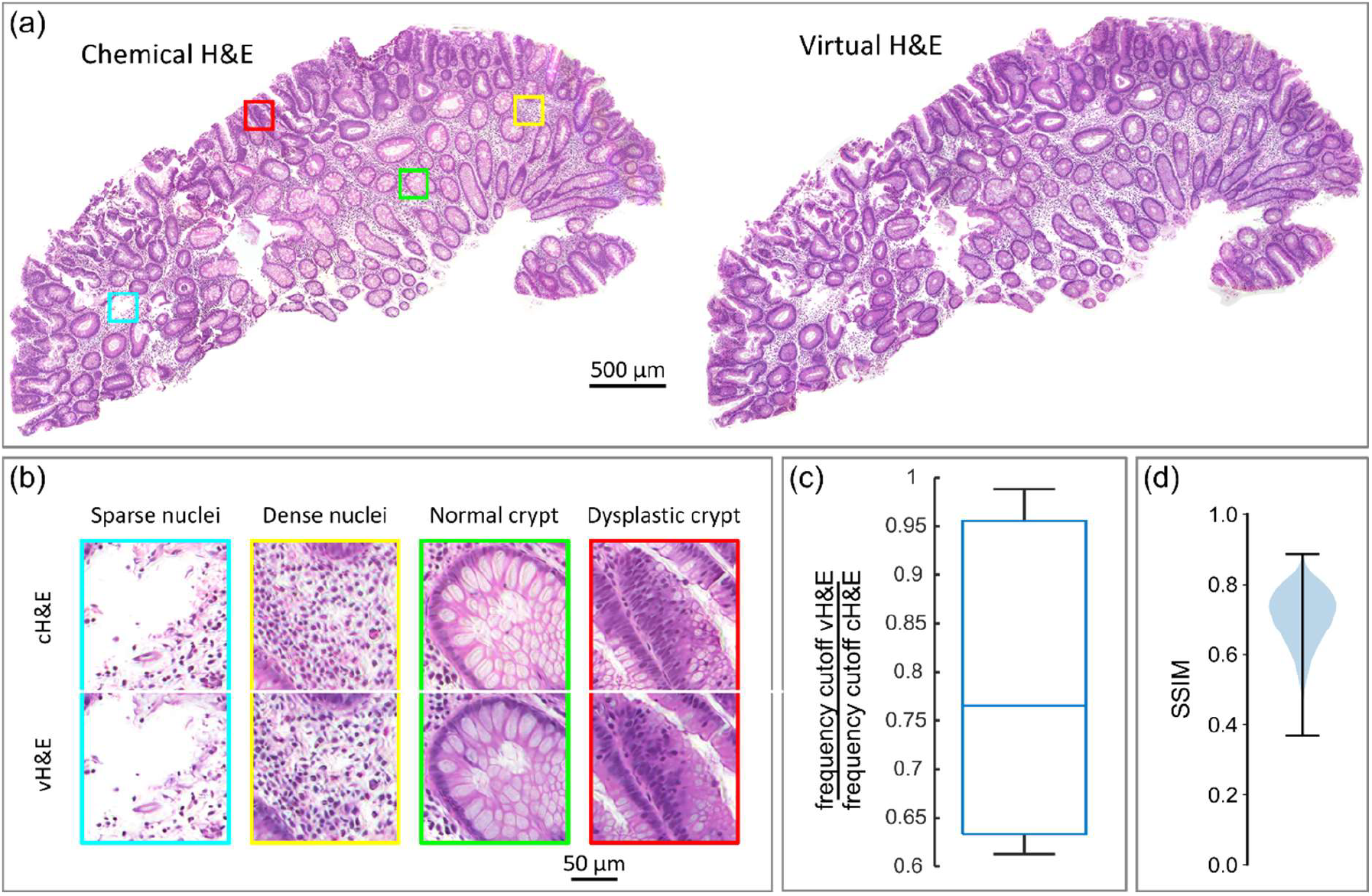
Comparison of chemically (c) H&E-stained FPM amplitude images and virtually (v) H&E-stained FPM phase images. (a) Representative cH&E and vH&E images of a colonic polyp section. (b) Magnified images of the boxed regions shown in panel (a), showing features in cH&E and corresponding vH&E image. (c) Box plot showing the relative cut of frequency for vH&E images compared to cH&E images for the images shown in (a). (d) Violin plot of structural similarity index for patches sampled from corresponding cH&E and vH&E images for six different polyp sections.

The similarity of cH&E and vH&E FPM images was quantified by computing the SSIM for matched images of the six polyp sections excluded from the training data (Fig. 3(d)). The mean SSIM over 3498 patches was 0.71 ± 0.08. The visual and quantitative similarity between chemically- and virtually-stained images is particularly striking considering the extensive sample processing (coverslip removal, staining and re-coverslipping) which took place between acquisition of image data for the unstained and chemically stained tissues. The results are comparable to those reported in other VS studies, such as Rivenson et. al. (Rivenson et al., 2019a) who obtained SSIM values of 0.81-0.89 for VS quantitative phase images of skin, kidney and liver tissue captured using a custom-built lensless holographic light microscope.

### 3.3 Comparison of WSI images of H&E-stained sections and VS-FPM images of unstained sections

As a final test of VS-FPM we compared images of chemically-stained polyp sections captured using a brightfield slide scanner to VS-FPM images of the same sections inferred from phase images captured prior to H&E staining. Aside from colour differences (discussed previously), when viewed at low magnification (Figure 4(a)) vH&E and cH&E results are visually similar, with the same tissue structure and large-scale features (including crypts and variations in cell number and density) apparent in both image data sets. However, the compound effects of differences between the FPM and the slide scanner systems and virtually and chemically-stained FPM images, mean that when viewed at higher magnification (Fig 4(b)) there are apparent differences between the images. These differences are particularly noticeable where nuclei are densely packed (such as in the lower right corner of Fig. 4(b)). The mean SSIM for VS-FPM and WSI results is 0.43 ± 0.07, similar to the mean SSIM for FPM images and WSIs of chemically-stained sections (0.51 ± 0.11); as noted previously, the mean SSIM for VS-FPM and FPM is relatively high (SSIM = 0.71 ± 0.08). PSNR and normalised RMSE values (Fig. S5) also indicate that VS-FPM and FPM images are more similar than cH&E FPM images and WSIs. Taken together, these results suggest that differences between vH&E images cH&E WSIs are due primarily to differences between the slide scanner and FPM; rather than reflecting a fundamental limitation of inferring vH&E from FPM phase images.

**Figure 4.**
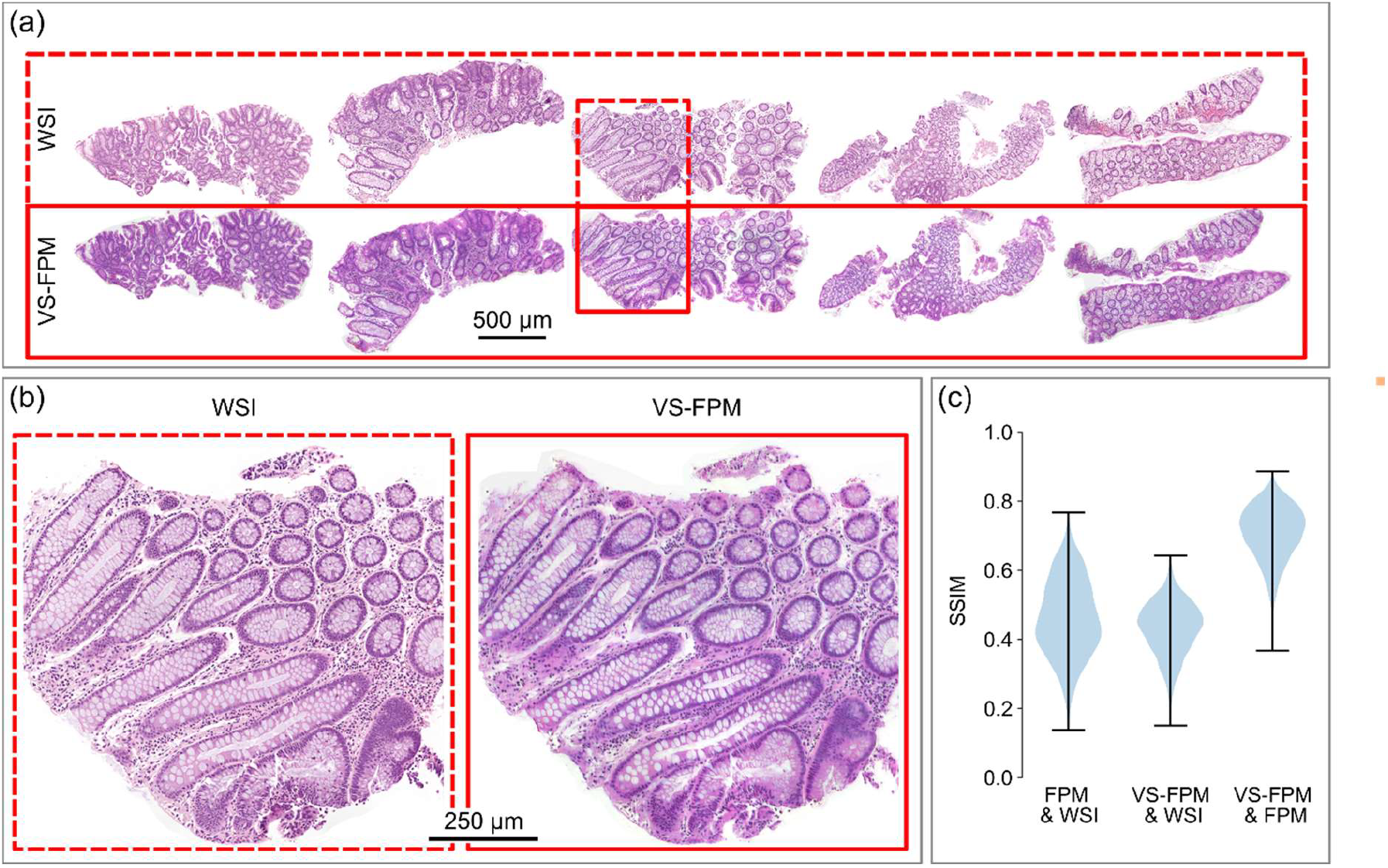
Comparison of WSI images of H&E-stained sections and VS-FPM images of unstained sections. (a) Low magnification images of brightfield WSI (top) and virtually-stained FPM phase images for five polyp sections. (b) Magnified images of a region of interest from one tissue section. (c) Violin plots showing SSIM values for permutations of H&E-FPM, WSI and VS-FPM image results. Mean values are SSIM_H&E FPM – WSI_ = 0.51, SSIM_VS-FPM – WSI_ = 0.43 and SSIM_H&E FPM – VS-FPM_ = 0.71

SSIM metrics are widely used to assess differences between images in radiology (Renieblas et al., 2017) and digital pathology (Hetz et al., 2024), however the values do not always correlate exactly with assessments made by human observers. Further, SSIM and other image similarity or difference metrics (Fig. S4&5) only provide a means to compare images, not assess their absolute fidelity. In the case of the present work our implicit and necessary assumption was, as the current gold standard for digital pathology, the brightfield WSIs from the slide scanner represented a ground truth. However studies have highlighted variations in images from (nominally similar) brightfield slide scanners which impact measurements of diagnostic features (Duenweg et al., 2023). To test the diagnostic utility of cH&E and vH&E FPM images, they were reviewed independently by two board-certified pathologists who were blinded to the diagnoses. In all 18 cases, both pathologists were easily and accurately able to distinguish normal from dysplastic tissue and derive correct pathological diagnoses.

## 4. Discussion and conclusions

We have shown that FPM phase images of unstained colonic polyps can be virtually stained using a conditional generative adversarial network to recover brightfield-like H&E images from which reliable and reproducible histological diagnoses can be made. FPM has several features which make it particularly attractive for virtual histological staining. Phase images recovered before and after histological staining significantly simplify the image registration process required to create paired images for network training. Previous work using phase imaging for VS (Rivenson et al., 2019a) used a multistep process which included training a separate NN for approximate phase to brightfield transformation to enable pixelwise registration. Decoupling the microscope field of view from the (diffraction-limited) spatial resolution of the objective enables entire tissue sections to be contained within a single field of view, reducing the need for sample scanning. Capture of image data using a low NA objective lens increases the depth of field of the microscope enabling digital post image capture refocusing (Claveau et al., 2020) to correct sample tilt and defocusing errors. A conventional transmitted light microscope can be converted into an FPM system by adding a low-cost LED array and microprocessor to synchronize illumination with the camera exposure, meaning the method can be readily adopted in clinical and research laboratories.

Empirically, FPM amplitude images of chemically-stained polyp sections were found to have a spatial resolution equivalent to images captured using a slide scanner with a 20x/0.75 objective lens. However, vH&E images were found to have spatial resolution approximately 20% lower than cH&E equivalents – likely due to image downsampling during inference. The use of high-definition GAN architectures (Wang et al., 2018) may offer a way to minimise this resolution loss and preserve fine features required for some diagnostic applications. Visual differences between FPM amplitude and WSI images were due primarily to colour variation and the development of improved colour calibration methods for FPM and VS-FPM is one way in which the VS-FPM results could be better matched to WSI data. However, even with conventional WSIs, a lack of colour standardisation remains an issue in digital pathology. Significant variations in colour have been reported even when imaging samples using nominally similar scanner technologies due to differences in specimen thickness, staining methods and scanner parameters (Inoue and Yagi, 2020).

As highlighted in the Introduction, VS methods are built from two core elements: a method for capturing images of unstained tissues and a computational workflow for translating label-free image information into histologically-stained images. VS-FPM can be further developed by advancing both of these. We observed significant differences between the sample phase images captured before and after chemical staining (see Fig. S1), due to absorption of light by H&E at the wavelength (530 nm) of the green LEDs in our FPM system. Other studies (Ban et al., 2018) have found that the effect of histological staining on phase images is negligible when using a light source at a wavelength range for which there is minimal absorption from the stain. As a result, we anticipate that modifying our setup to include a longer wavelength red LED (∼730 nm) would enable the virtual staining network to be trained using data (phase and amplitude images) from histologically-stained tissue alone. FPM hardware can be readily modified to capture more information by detecting perturbations to other properties of the microscopic vector light field as it propagates through the sample. For example, the addition of polarizing elements enables visualization of the birefringence properties of biological specimens (Song et al., 2021). High-resolution FPM systems can recover amplitude and phase images with a synthetic NA > 1 (Ou et al., 2015), enabling high-resolution diagnostic imaging applications such as karyotyping (K. Zhang et al., 2022). As we have demonstrated, subcellular features are visible in FPM phase images due to refractive index differences and we anticipate that a high-resolution implementation of VS-FPM would enable label-free VS for further applications in histopathology and cytopathology. In addition to supervised GANs, such as pix2pix, researchers have successfully employed cycle-consistent GANs (Zhu et al., 2017) - which do not require pixelwise registration of training data. A variety of other DNN architectures have been investigated (Wang et al., 2024) and the optimal method for generating a given histological stain from a given label-free imaging modality remains an open question.

Having demonstrated VS-FPM for a specific use case an obvious question is: can it generalize to other tissue types and stains? Measurements have shown that refractive index of the cell nucleus is lower than that of the cytoplasm (Steelman et al., 2017), which explains why nuclei are clearly visible in FPM phase images (Fig. S6). The characteristic ellipsoidal shape of the nuclei, visible against the rest of the tissue, provides an explanation for how a cGAN is able to apply a virtual haematoxylin stain. Previous studies have demonstrated that quantitative phase images of liver and kidney can be virtually stained using Masson’s trichrome and Jones’ stains and we anticipate that VS-FPM is likely to be effective for virtually staining tissues with these and other special stains which highlight tissue structure. Further, stain-to-stain translation methods (Burlingame et al., 2020;R.Zhang et al., 2022) offer a way to perform virtual IHC staining of unstained tissues via reconstructed vH&E images.

Obviating the need for chemical staining has clear benefits for analysis of FFPE tissue sections, however VS methods also have wider applications in histopathology. Our previous work (Claveau et al., 2020) has demonstrated that numerical propagation of the complex field recovered by FPM enables post capture refocusing and reconstruction of extended depth of field images over an axial range corresponding to the depth of field of the objective lens (approximately 20 μm for the system used in this study). This suggests VS-FPM may be suitable for analysing thicker specimens such as cell smears in cytopathology and sections of fresh, unprocessed tissue in intraoperative histopathology. A particular advantage of FPM, compared to other label free imaging techniques, is the relative simplicity of the hardware. In principle, any transmitted light microscope can be configured for VS-FPM by modifying the illumination pathway to incorporate an LED array which is synchronized to the microscope camera. This means that VS-FPM can be adapted for different spatial resolution requirements to suit a range of different applications in histopathology and cytopathology. The relatively low cost and wide availability of LED arrays and microcontrollers means there is a low barrier to adoption of the technology in resource constrained settings, particularly as recovery of the microscopic pupil function enables correction of imaging aberrations associated with low cost imaging hardware (Collins et al., 2020; Diederich et al., 2020).

## CRediT authorship contribution statement

**Christopher Bendkowski**: Data curation, Formal analysis, Investigation, Methodology, Software, Vizualization, Writing – original draft, Writing – review & editing. **Adam Levine**: Resources, Validation, Writing – review & editing. **Manuel Rodriquez-Justo**: Resources, Validation, Writing – review & editing. **Laurence Lovat**: Funding acquisition, Writing – review & editing. **Marco Novelli**: Conceptualization, Resources, Validation, Writing – review & editing. **Michael Shaw**: Conceptualization, Data curation, Formal analysis, Funding acquisition, Investigation, Methodology, Project Administration, Resources, Software, Supervision, Validation, Vizualization, Writing – original draft, Writing – review & editing

## Acknowledgements

The research was funded by a proof-of-concept award from Wellcome/EPSRC Centre for Interventional and Surgical Science (Wellcome Trust) 203145Z/16/Z and the NIHR UCLH Biomedical Research Centre. APL was funded by The Pathological Society of Great Britian and Ireland.

## Data availability

The image data presented in this article are available at https://doi.org/10.6084/m9.figshare.29086454.

