## Supplementary material for "VS-FPM: large-format, label-free virtual histopathology microscopy": Suplementary information

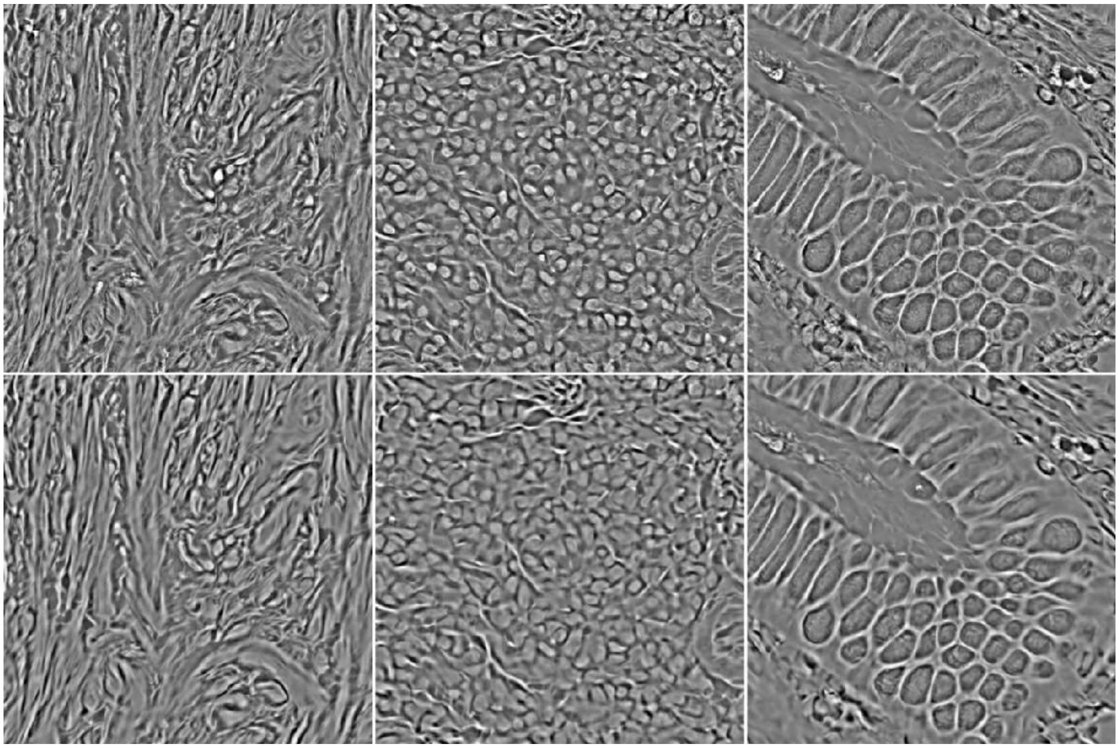

**Figure S1. Representative FPM phase images of the same tissue section before (top) and after (bottom) H&E staining**.

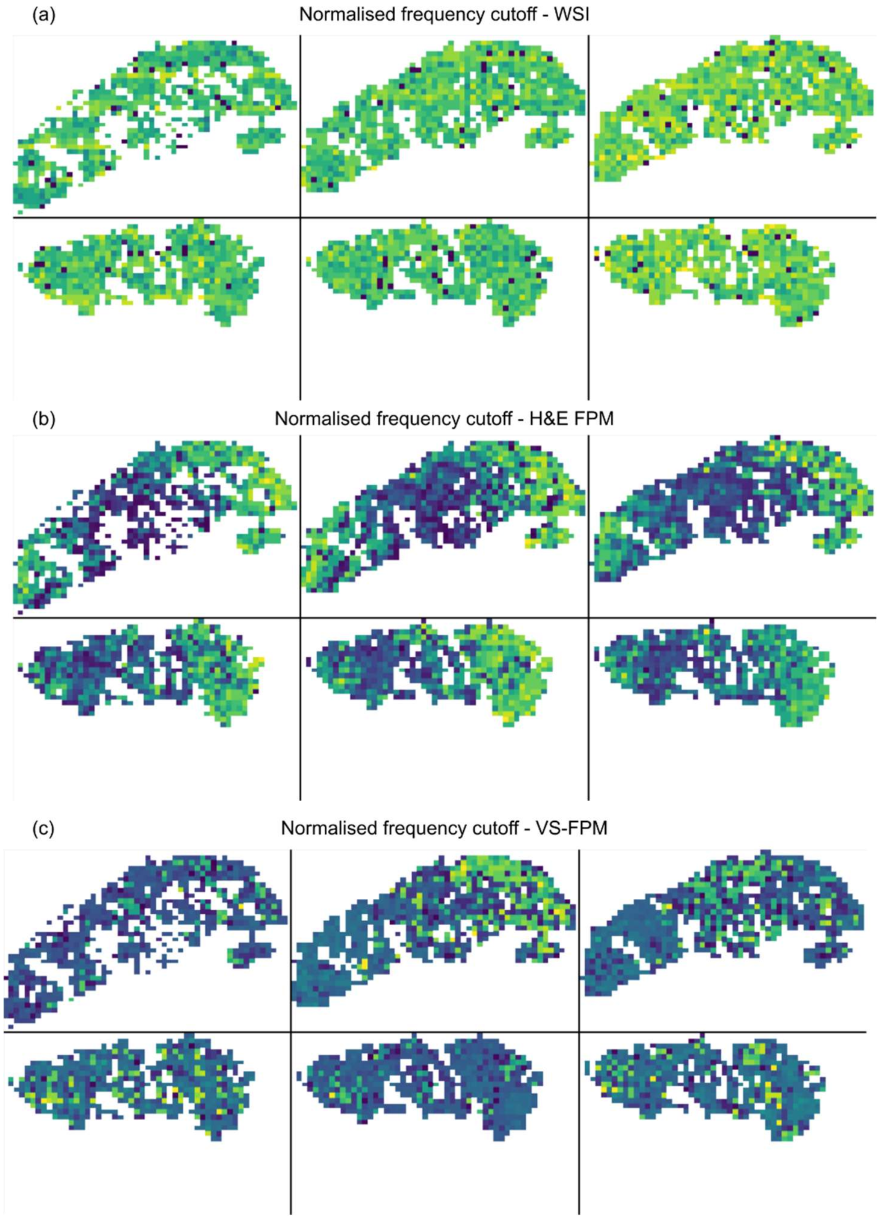


**Figure S2. Heatmaps showing spatial variation in normalized spatial frequency cutoff.** (a) WSI and (b) FPM amplitude images of six H&E-stained polyp tissue sections, and (c) virtually H&E-stained FPM phase images for the same sections prior to chemical staining. Heatmaps min-max normalized for each modality.

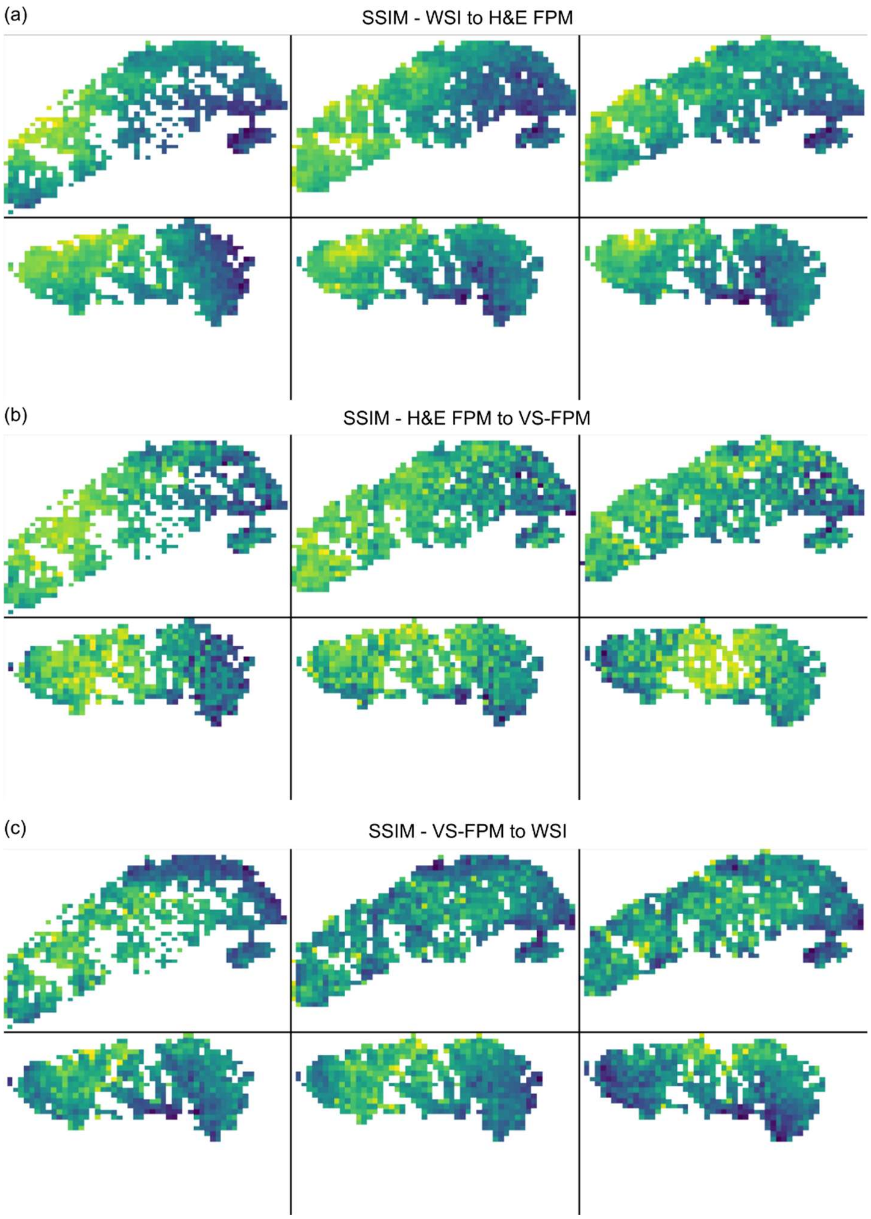


**Figure S3. Heatmaps showing spatial variation in structural similarity of paired images for six polyp sections.** (a) SSIM heatmaps for WSI against FPM amplitude images for H&E stained sections. (b) SSIM heatmaps for FPM amplitude image of H&E stained tissue against virtually H&E stained FPM phase images. (c) SSIM heatmaps for WSI against virtually H&E stained FPM phase images. Heatmaps min-max normalized for each modality pairing.

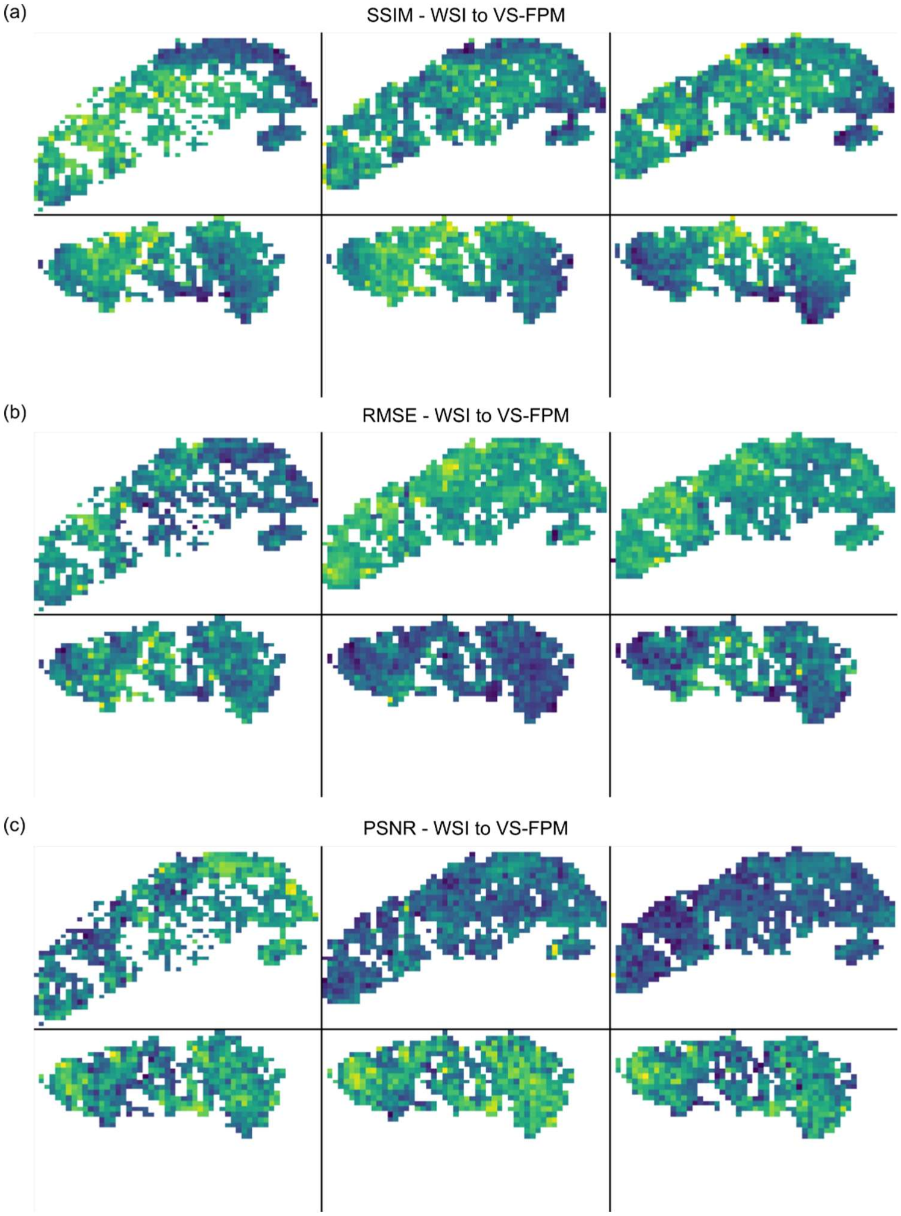


**Figure S4. Spatial variation in (a) SSIM, (b) RMSE and (c) PSNR for WSIs H&E-stained polyp sections and VS-FPM images of the same sections captured prior to chemical staining**. Heatmaps min-max normalized for each metric.

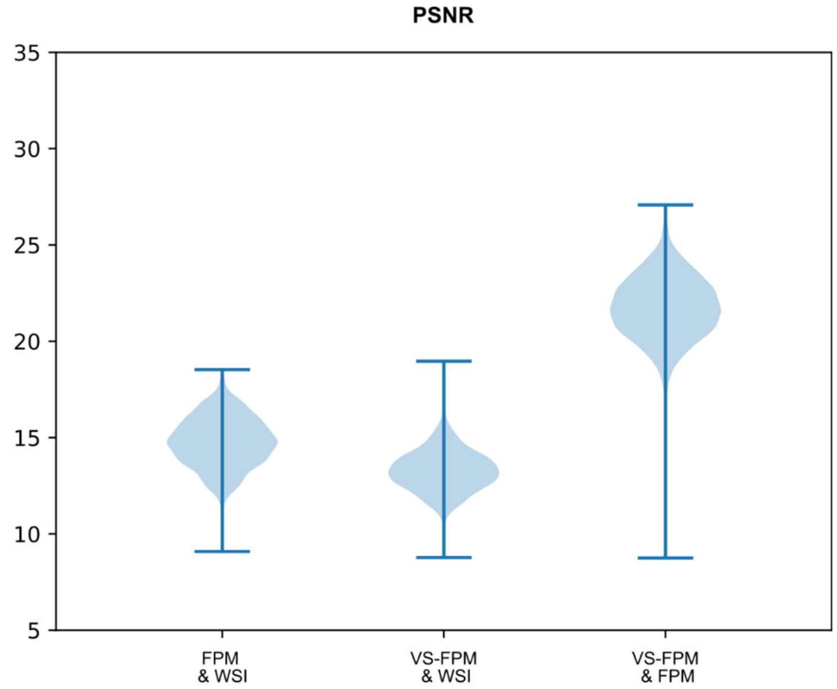


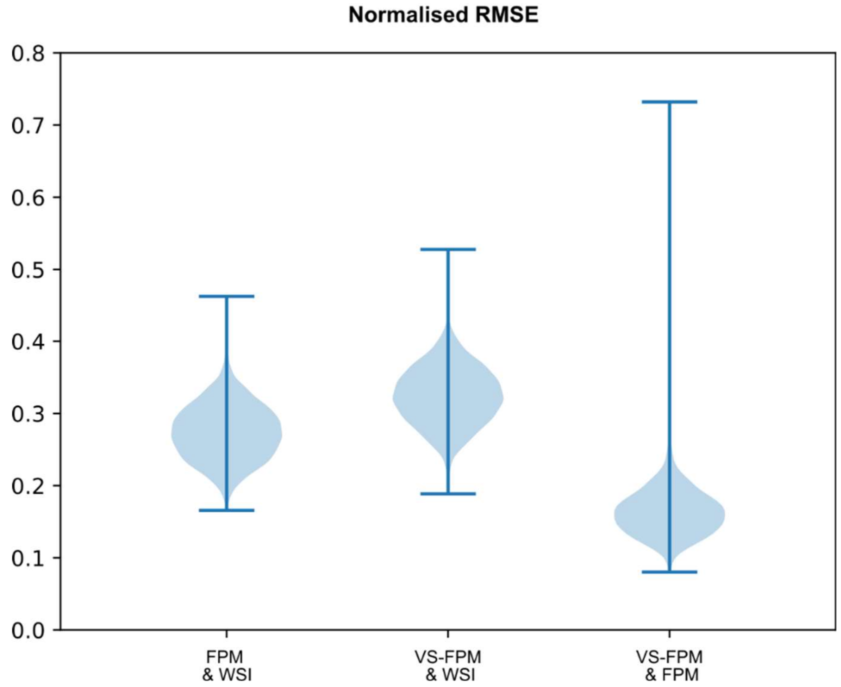


**Figure S5. Peak signal-to-noise (PSNR) and normalized root mean square error (RMSE) image difference metrics for WSIs and FPM amplitude images of H&E-stained sections, VS-FPM images and WSIs of virtually and chemically stained sections, and VS-FPM and FPM amplitude images of virtually and chemically stained sections.** Data corresponds to 3498 image patches from six different polyp sections.

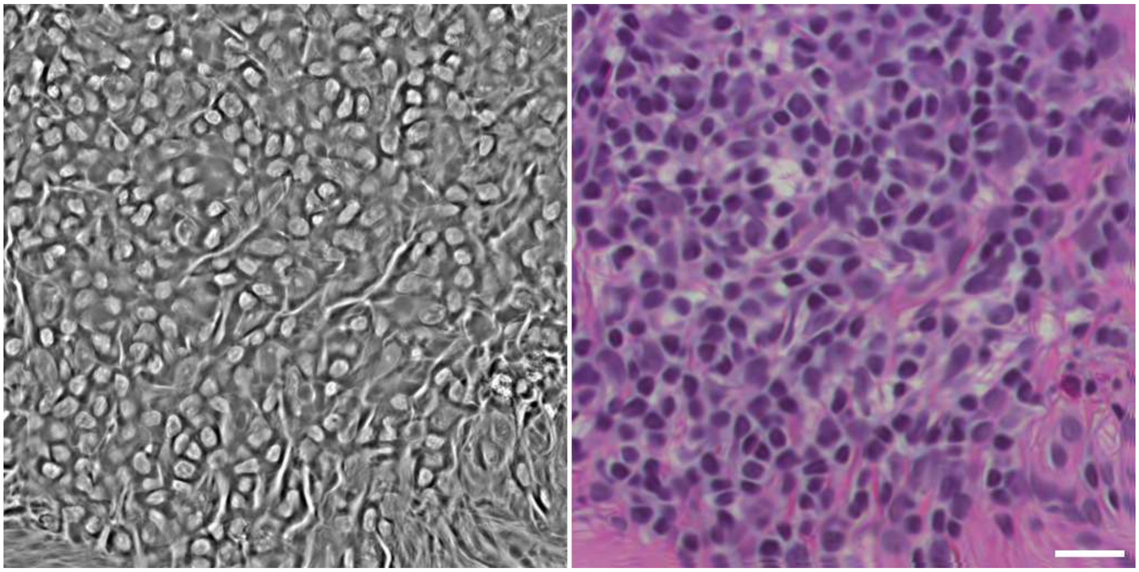

**Figure S6. Representative FPM phase image (left) of an unstained colonic polyp and corresponding FPM amplitude image (right) after chemical H&E staining.** In the phase image cell nuclei are visible as bright quasi-ellipsoidal blobs on a grey background. Scale bar 20 µm.
